# Whole genome analysis in *APOE4* homozygotes identifies the *DAB1-RELN* pathway in Alzheimer’s disease pathogenesis

**DOI:** 10.1101/2022.04.28.22274418

**Authors:** Matthew Bracher-Smith, Ganna Leonenko, Emily Baker, Karen Crawford, Andrew C. Graham, Dervis A. Salih, Brian W. Howell, John Hardy, Valentina Escott-Price

**Affiliations:** MRC Centre for Neuropsychiatric Genetics and Genomics, Division of Psychological Medicine & Clinical Neurosciences, Cardiff University, UK; Dementia Research Institute, Cardiff University, UK; Dementia Research Institute, University College London, UK; Neuroscience and Physiology, State University of New York, USA

**Author notes:** Corresponding Authors* Valentina Escott-Price, Address: Hadyn Ellis Building, Cardiff University, Maindy Road, Cardiff, CF24 4HQ, UK, John Hardy, Address: UCL Cruciform Building, Gower Street, London, WC1E 6BT, UK.

## Abstract

The *APOE*-ε4 allele is known to predispose to amyloid deposition and consequently is strongly associated with Alzheimer’s disease (AD) risk. There is debate as to whether the *APOE* gene accounts for all genetic variation of the *APOE* locus. Another question which remains is whether *APOE*-ε4 carriers have other genetic factors influencing the progression of amyloid positive individuals to AD. We conducted a genome-wide association study in a sample of 5,390 *APOE*-ε4 homozygous (ε4ε4) individuals (288 cases and 5,102 controls) aged 65 or over in the UK Biobank. We found no significant associations of SNPs in the *APOE* locus with AD in the sample of ε4ε4 individuals. However, we identified a novel genome-wide significant locus associated to AD, mapping to *DAB1* (rs112437613, OR=2.28, CI=1.73-3.01, *p*=5.4×10^−9^). This identification of *DAB1* led us to investigate other components of the *DAB1-RELN* pathway for association. Analysis of the *DAB1-RELN* pathway indicated that the pathway itself was associated with AD, therefore suggesting an epistatic interaction between the *APOE* locus and the *DAB1-RELN* pathway.

## Introduction

Genome wide association studies (GWAS) have led to the identification of many genetic loci influencing the risk of dementia [1]. However, none of these approach the importance of the *APOE* locus [2] where the *APOE-*ε4 allele has a frequency of ∼15% in controls and has a risk ratio of >3 in cases. Other loci with allele frequencies of >1% have risk ratios of <1.4. Recent studies have shown that the *APOE* genotype is almost solely responsible for amyloid deposition whereas other components of Alzheimer’s disease (AD) genetic risk contribute to the occurrence of dementia in the context of amyloid deposition [3]. Furthermore, neuropathologic studies have shown that clinical diagnoses in Alzheimer series had a diagnostic accuracy of around 80%: this accuracy is implied by analyses comparing the large clinical GWAS with the smaller neuropathologic GWAS, leading to the concern that these larger GWAS are contaminated by other diagnoses. This concern is heightened by the reports of loci for frontotemporal dementia in case series labelled as Alzheimer’s disease in the most recent GWAS for the disorder [4].

With this background, we have undertaken an AD GWAS in individuals who are *APOE-*ε4 homozygotes for three reasons. First, because in this group diagnostic accuracy is very high; second, to assess whether in this context there is additional genetic risk at the *APOE* locus; and third, to assess which previously reported loci are replicated in these cases and whether there are any novel loci we can identify which are dependent on *APOE-*ε4 homozygosity. This study was possible in the UK Biobank [5] because it has a very large cohort, with a sufficient number (for statistical analyses) of *APOE-*ε4 homozygotes, where many participants are now reaching the age where they are at risk.

Here we report that the *APOE* allele alone accounts for the AD risk in the LD block on chromosome 19 in the European population. Furthermore, in *APOE-*ε4 homozygotes, we identify AD risk associated with the *DAB1* gene that encodes a synapse regulatory protein. Subsequent analyses revealed a gene set association with the *DAB-RELN* pathway.

## Methods

### Phenotypes

Individuals from the UK Biobank were considered if they self-reported as white British and were of similar genetic ancestry by principal component analysis (UK Biobank field 22006), were unrelated (kinship coefficient < 0.04) and if they had not withdrawn consent to participate under UK Biobank. Participants were further excluded if they showed excessive missingness or sex chromosome aneuploidy, were outliers for heterozygosity, had mismatching self-reported and inferred sex from genotyping data, and had over 10 putative third-degree relatives. AD definition was derived using ICD-10 codes in hospital and death records. Individuals were coded as cases where dementia in Alzheimer’s disease (ICD-10 code F00) or Alzheimer’s disease (code G30) were present. Controls were defined as those without F00, G30, vascular dementia (F01), dementia in other diseases (F02) and unspecified dementia (F03). *APOE* status was assigned to each individual, as defined by SNPs rs7412 and rs429358 which are both present on the Affrymetrix Axiom genotyping array used. After quality control and restriction to *APOE-*ε4 homozygous individuals aged 65 or over, 288 cases and 5,102 controls were included in analysis.

### Genetic quality control

The UK Biobank genetic data from the haplotype reference consortium (HRC), imputed by the UK Biobank [6], was restricted to biallelic SNPs (minor allele frequency > 0.05) with Hardy-Weinberg equilibrium > 10^−6^, INFO>0.4 and posterior probability>0.4. After quality control, 5,349,830 SNPs were included in analysis.

### Analysis

Association analysis was conducted in PLINK2 [7] on UK Biobank dosage data using most recently recorded age, sex and the first 15 principal components (field 22009) as covariates. The significant findings (with the logistic regression) were further tested with Cox proportional-hazards regression (while controlling for the covariates) where the censoring occurred when a participant reported AD, allowing for the fact that some individuals have not reached the age at onset and may develop the disease given time.

The enrichment analysis of significant SNPs (at 5% significance level) or for SNPs showing the same direction of the effect (assuming that the chance to have the same direction of effect is 50%) was performed with binom.test() function in R.

The power calculations were performed with qnorm() function in *R*-statistical package at nominal 5% significance level (unless specified otherwise), where Z-score was estimated as log(OR)/var with the log(OR) as reported in the GWAS. In the Wightman et al. study [4], the largest OR was selected from the reported ORs in the list of contributing studies. The variance estimated as the inverse variance, with allele frequencies in cases and controls (corresponding to the SNP OR), and the sample size as in our study (*N* cases = 288, *N* controls = 5,102). Plots of regional associations were created using LocusZoom [8].

Epistasis was defined as deviation from joint two SNPs linear effects in the logistic regression model (known as statistical interaction). Significance of the interaction term was assessed using --epistasis option PLINK [7], accounting for the same covariates as above. The interaction plots were produced using matplotlib in python [9].

Gene-based analysis was run by MAGMA using FUMA v1.3.7 [10, 11]. Competitive setting of MAGMA was also used to test the candidate pathways for the enrichment of AD significant genes as compared to the rest of the genome.

### DAB1-RELN pathway analysis

The canonical Reelin-Dab1 signalling pathway has been studied extensively in mouse neurons and brain [12]. For analysis, we divided the pathway into three sections: a) the receptor complex, (Reelin, the receptors ApoER2, VLDLR, the adaptor protein DAB1, and the tyrosine kinases SRC, FYN and YES) [13–16], b) branch 1 that regulates N-cadherin (CRK, CRKL, C3G, RAP1, P120 catenin, N-cadherin) [17–19] and c) branch 2 that is involved in microtubule-associated protein tau (MAPT) phosphorylation (PI3K, PDK, AKT, GSK3, STK25) [20–22]. We converted these mouse proteins to the homologous human genes with the BioConductor function in R and the NCBI database (www.ncbi.nlm.nih.gov/) yielding: a) *RELN, LRP8, VLDLR, DAB1, SRC, FYN, YES1, b) CRK, CRKL, RAPGEF1, RAP1A, CTNND, CDH2, c) PIK3CA, PDK1, PDK2, GSK3B, AKT1, STK25*.

## Results

We present the results in the following order: (a) analysis of the *APOE* locus, (b) analysis of other previously reported GWAS in these cases, (c) identification of the *DAB1* locus as a genome wide for disease, (d) assessment of other loci in the same *DAB1-RELN* pathway.

### *APOE* locus

No suggestive variants were identified in the *APOE* gene or surrounding region (chromosome 19: 44.5-46.5 Mb, as defined previously [23]) with the lowest *p*-value at 0.003 within 1Mb of the *APOE* gene (Supplementary Figure S1) in *APOE-*ε4 homozygotes.

### Other GWAS Hits

Loci previously reported as GWAS for association with Alzheimer’s disease status did not show a strong replication in the current analysis of *APOE*-ε4 homozygotes only (Supplemental Table S1). Though the power to detect the GWAS-reported effect sizes in this sample is not sufficient (see last column of Supplemental Table S1), four loci in *CD33* (*p*=0.004), *IQCK* (*p*=0.009), *LILRB2* (*p*=0.005) and *SORL1* (*p*=0.007, MAF=0.04) had the strongest evidence for association in the current analysis and a consistent direction of effect between the current and previous GWAS. Weaker but nominally significant associations with the consistent direction of the effect were also observed in the *APH1B* (*p* = 0.024), *BIN1* (*p*=0.011), *SEC61G* (p=0.015) and *SNX1* (p=0.048) genes. In total, eight out of 77 SNPs (previously reported as genome-wide significant and available in our study), replicated at significance level with the same direction of association, which is statistically greater than chance (p=0.038). In addition, 53/77 (69%) SNPs have same direction of effect in the current analysis and previous GWAS which is greater than expected by chance (p = 0.001).

### Identification of DAB1 as a locus

Multiple novel genome-wide significant intronic SNPs were present in *DAB1* (lead SNP: rs112437613, OR=2.28, CI=1.73-3.01, *p*=5.36×10^−9^; Figure 1 and Supplemental Figure S2, Table 1). The minor allele T was associated with disease risk (MAF=6% in non-AD and 12% in ADε4ε4-participants of the UK Biobank). To allow for the fact that some individuals might not have reached the age at onset, we fit a survival regression model (adjusting for PCs and sex). The result remained highly significant (Hazard Ratio=2.27, CI=1.75-2.95, p= 7.8×10^−10^). The Kaplan-Meier graph (Figure 2) demonstrates that probability of getting the disease (y-axis) earlier (x-axis) is higher as the number of the risk alleles of rs112437613 SNP increases.

**Table 1:** Novel genome-wide significant SNPs in *DAB1*.

| CHR | BP | SNP | Closest gene | Current Analysis |  |  |  |  | Previous LOAD GWAS [65] |  |  |
| --- | --- | --- | --- | --- | --- | --- | --- | --- | --- | --- | --- |
|  |  |  |  | Effect/Alt | Freq | OR | SE | p-value | Effect/Alt | OR | p-value |
| 1 | 57625932 | rs17541203 | *DAB1* | C/T | 0.07 | 2.19 | 0.14 | 2.4x10<sup>-8</sup> | C/T | 0.98 | 4.8x10<sup>-1</sup> |
| 1 | 57643271 | rs197111 | *DAB1* | T/C | 0.07 | 2.16 | 0.14 | 3.1x10<sup>-8</sup> | T/C | 0.98 | 4.1x10<sup>-1</sup> |
| 1 | 57646630 | rs78921149 | *DAB1* | T/C | 0.07 | 2.22 | 0.14 | 1.4x10<sup>-8</sup> | T/C | 0.98 | 4.2x10<sup>-1</sup> |
| 1 | 57647715 | rs112437613 | *DAB1* | T/C | 0.07 | 2.28 | 0.14 | 5.4x10<sup>-9</sup> | T/C | 0.99 | 5.0x10<sup>-1</sup> |
| 1 | 57648856 | rs17115257 | *DAB1* | G/T | 0.08 | 2.12 | 0.13 | 1.0x10<sup>-8</sup> | G/T | 0.99 | 6.5x10<sup>-1</sup> |
| 1 | 57650410 | rs58359668 | *DAB1* | T/C | 0.08 | 2.12 | 0.13 | 1.1x10<sup>-8</sup> | T/C | 0.99 | 6.5x10<sup>-1</sup> |

CHR –chromosome; BP –base-pair position in build37; SNP –single nucleotide polymorphism, closest gene –genes were annotated with assembly hg19; effect/non-effect –effect and non-effect alleles; freq- frequency of reference allele in the UK Biobank *APOE*-ε4 homozygotes individuals; OR, SE, p-value –odds ratio, standard error and p-value of the current and previous reported AD GWAS association studies; GWAS –reference of the corresponding GWAS study

**Figure 1:**
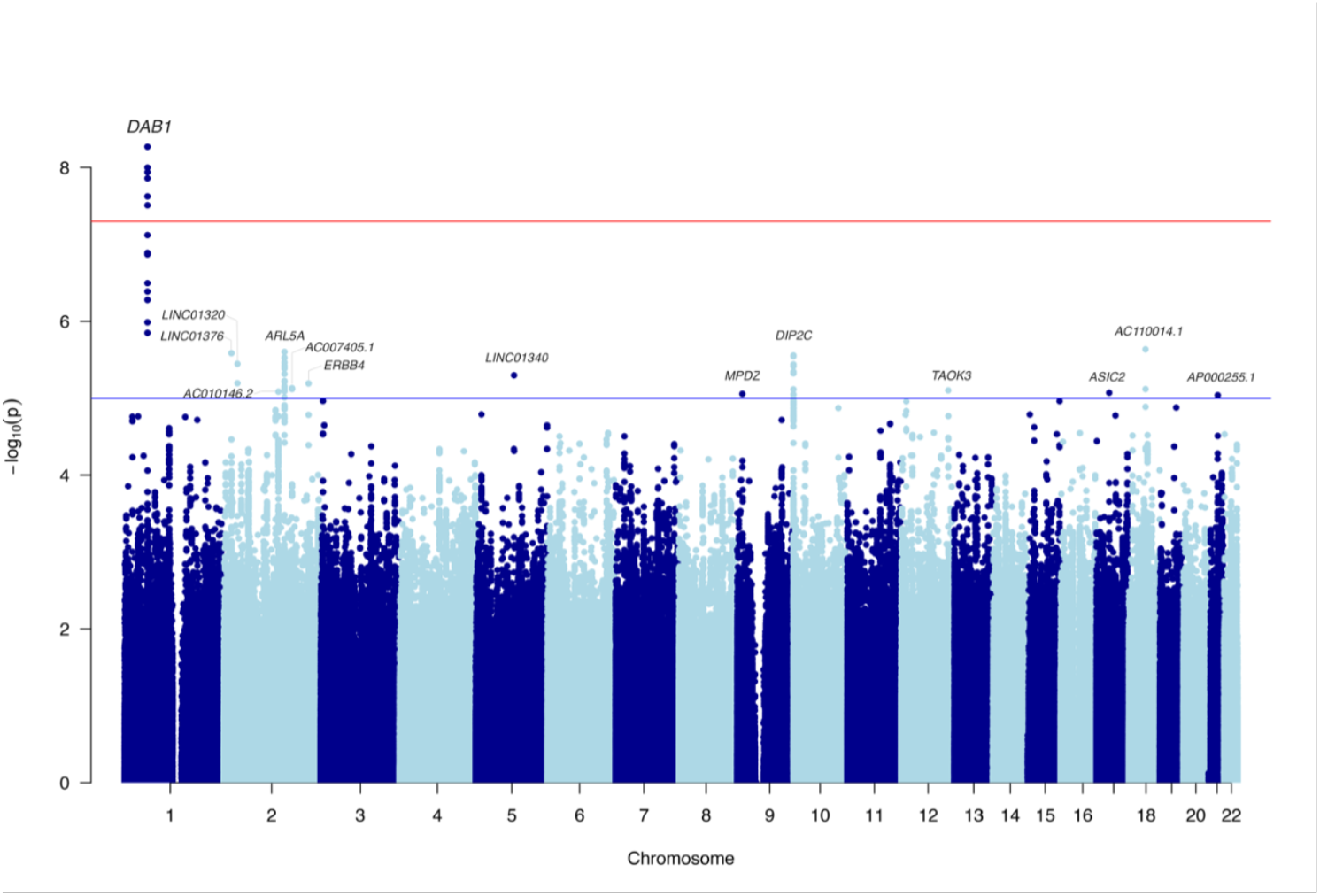
Manhattan plot for the genome-wide association study in *APOE*-ε4 homozygotes with SNP MAF > 5%.

**Figure 2:**
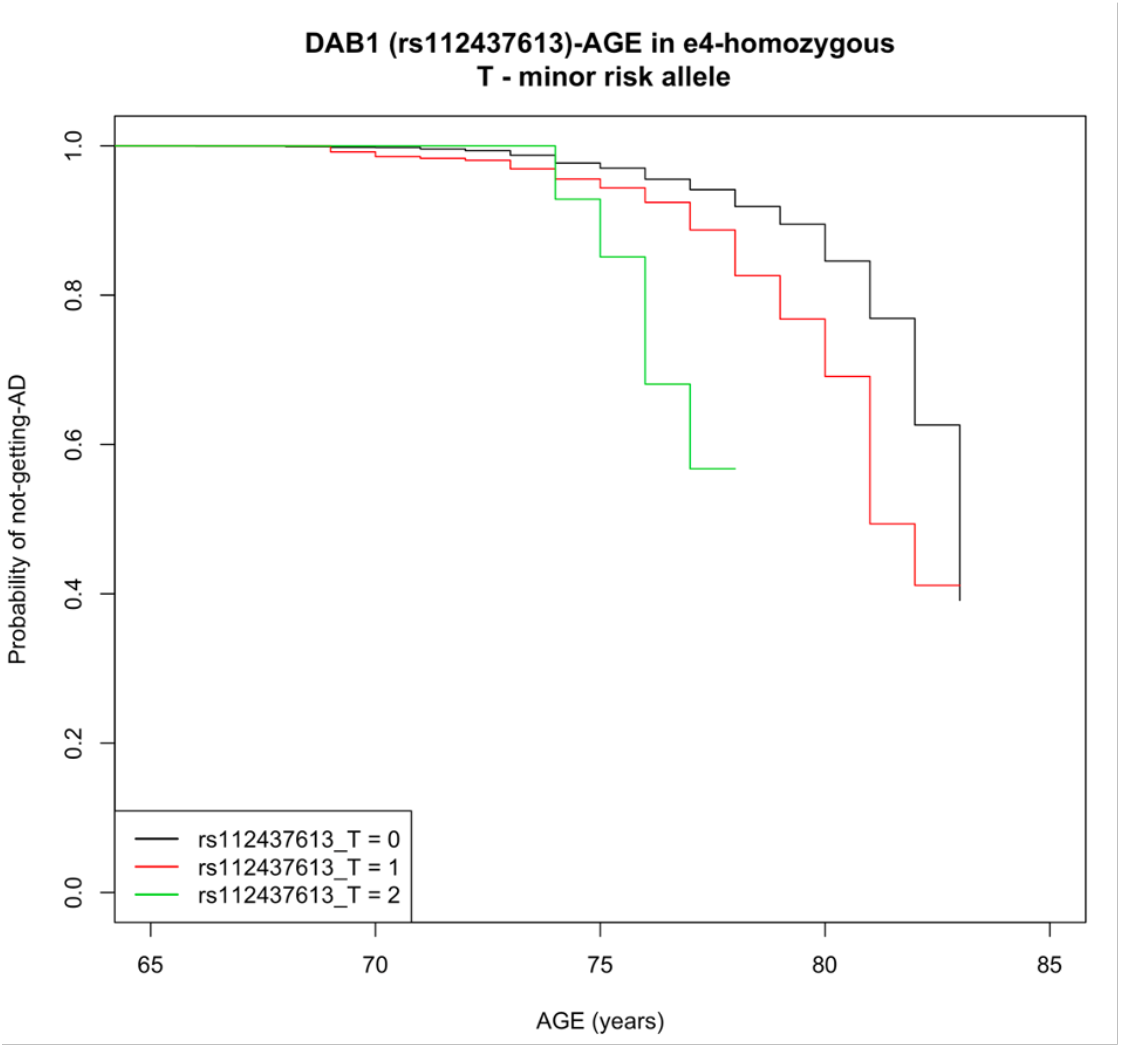
The cumulative risk of AD among *APOE*-ε4 homozygous of the UK Biobank participants, who carry 0, 1 or 2 risk alleles T the lead SNP (rs112437613) in *DAB1*.

**Figure 3.**
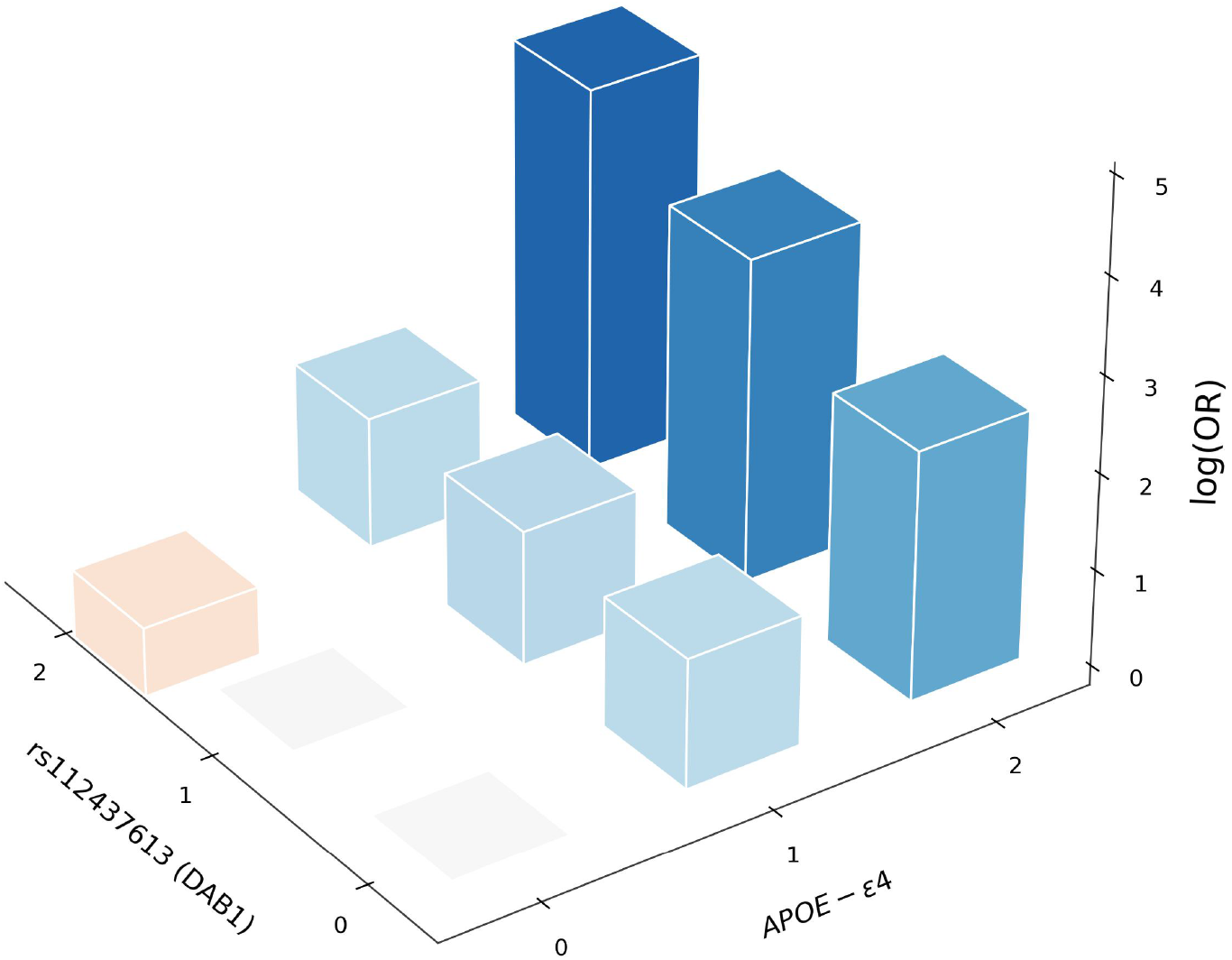
Epistatic effect between *APOE*-ε4 and rs112437613 (*DAB1*) in the whole sample of the UK Biobank aged 65+ (N=229,748). All log(odds ratio) values are with respect to the baseline homozygote with no counted alleles at both loc. Orange/red bars have negative values. All odds ratios are adjusted for age, sex and principal components.

**Figure 4.**
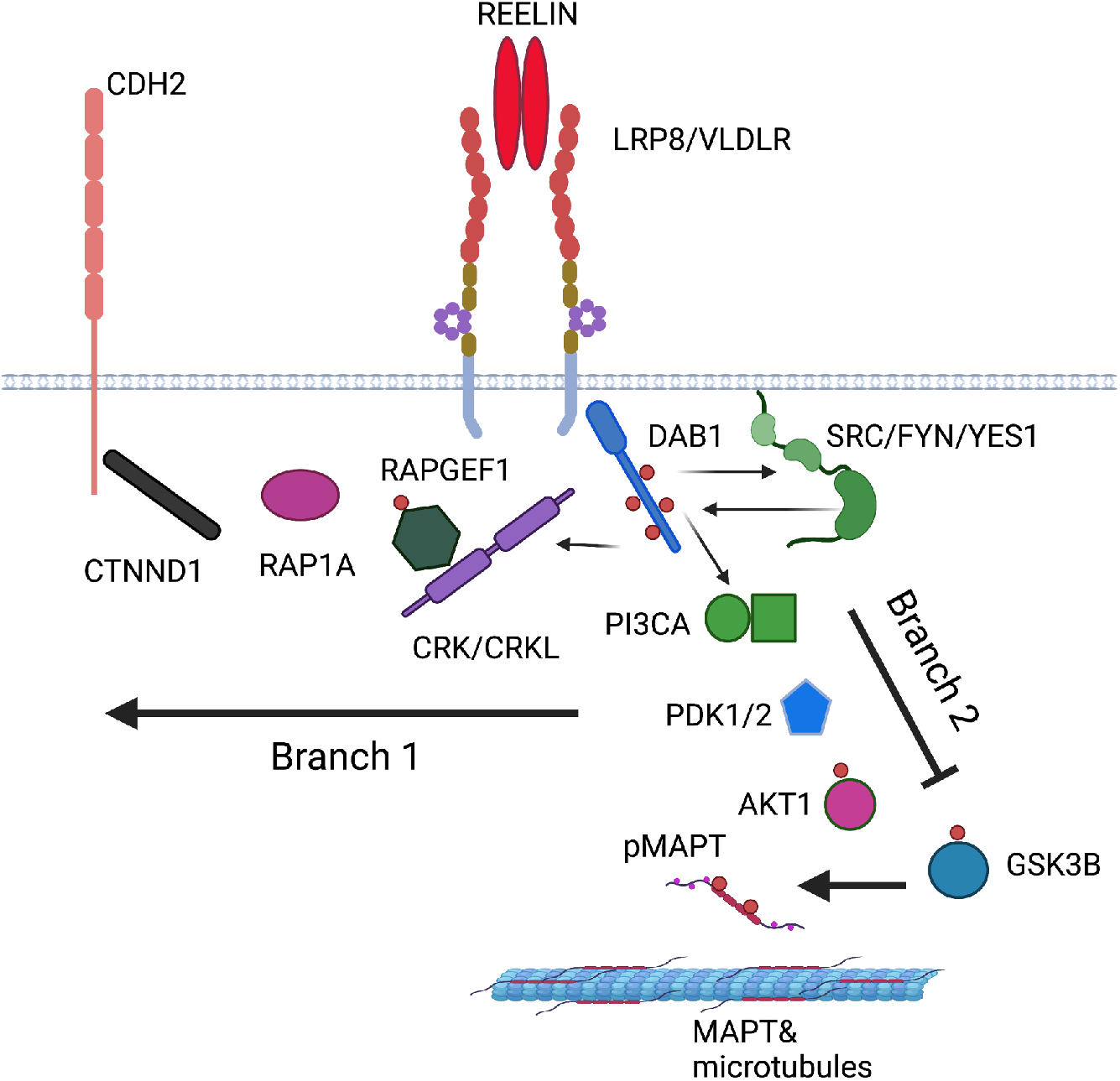
REELIN-DAB1 signalling pathway based on studies in mouse neurons and brain (human protein names are shown, created with BioRender.com, see BioRender’s Academic License publication licence in the Supplemental material). The pathway branches downstream of the signalling complex. Branch 1 regulates the cell surface expression of CDH2 (N-cadherin) and branch 2 regulates MAPT phosphorylation.

The frequency of this allele is reported 4%-7% in European population cohorts (1000Genomes, TOPMED, GnomAD, Estonian, ALSPAC-UK, TWINSUK, Northern Sweden, see https://genome.ucsc.edu). However, this SNP (and others in LD with it) did not show even a nominal association to AD in recent GWAS that did not preselect for the ε4ε4 genotype: e.g. a study of 21,982 cases and 41,944 controls the p-values were p∼0.5 (see Table 1) [24]. Indeed, in a case/control sample (without screening for the *APOE*-ε4 status), the effect size of this SNP would be OR=1.016, as the proportion of cases, with both T allele of rs112437613 and ε4ε4, is 0.016 (=MAF(ε4)^2^*MAF(rs112437613 in ε4ε4) = 0.36^2^*0.12, where 0.36 is the ε4 allele frequency in cases [25], and, similarly, of controls is 0.001. Therefore, the frequencies of the T allele in the overall sample are expected to be 0.061 in cases and 0.06 in controls, and consequently, the power to detect it with the sample size of the [24] study is close to 0 (∼3×10^−7^).

This observation led us to test for an epistatic effect in the whole sample of the UK Biobank aged 65+ (N=229,748). There was indeed significant epistasis between the two loci (p=1.5×10^−5^), whereas the effect of the T allele (rs112437613) was positive (OR=1.16, SE=0.11), but only nominally significant (p=0.021), providing evidence for cooperation between these two loci. The risk allele frequencies in this locus depending on *APOE* and AD status are shown in Table S2 and the risk of AD, depending on the genotypes at the two loci, is shown in Figure 5. The figure and table clearly show a statistical epistatic effect, where the disease risk is only visible in people with ε4ε4 genotypes.

### Candidate analysis of other loci in the Reelin-DAB1 pathway

*Dab1* encodes a cytoplasmic signalling adaptor that is predominantly expressed in neurons where it acts downstream of the extracellular ligand Reelin to regulate brain lamination during development [26–29]. Since Reelin-DAB1 signalling also performs an important role in the adult brain by promoting excitatory synapse maturation [30, 31] and modulating synaptic plasticity, learning and memory [32–35], we explicitly looked at the SNPs associations in the *RELN* gene (chr7:103,112,231-103,629,963). This gene is comprised of 2002 SNPs and the most significant SNP was rs171331137 (chr7:103479651) with OR=1.51 (SE=0.11), p=2.4×10^−4^ (Supplemental Figure S3). Similar to *DAB1*, we tested this SNP for interaction with *APOE*-ε4 in the whole UK Biobank sample. The interaction term was not significant (p=0.24), however the pattern of AD risk based on the pair of these markers was similar to *DAB1* (Supplemental Figure S4).

The Reelin ligand and DAB1 adaptor proteins are bridged by two partially redundant transmembrane receptors APOER2 (LRP8) and VLDLR [15, 16]. Reelin binding to its receptors recruits DAB1 to their cytoplasmic tails activating the SRC family kinases, SRC, FYN and YES [13, 14, 36]. This leads to the increased tyrosine phosphorylation of DAB1 and the recruitment of additional signalling adaptor proteins that activate two key branches of the pathway (Figure 6). One branch is initiated by the binding of CRK and CRKL to phospho-DAB1, leading to the phosphorylation of C3G (*RAPGEF1*) and activation of RAP1 (*RAP1A*) [17, 18]. This leads to the upregulation of N-Cadherin (*CDH2*) cell-surface expression through engagement with p120 catenin (*CTNND*) [19]. A second branch is regulated by the binding of phosphatidylinostiol 3-kinase (*PIK3KA*) to DAB1 leading to the activation of PDK (*PDK1, PKD2*) and AKT (*AKT1*) ultimately suppressing the activity of the MAPT kinase GSK3 [20]. In mouse, deficiency of DAB1 has been shown to augment tau-phosphorylation and Stk25 has been implicated in this process [21, 22]. Since the signalling complex and the downstream pathways have potential significance in the development of AD, we tested their associated genes for enrichment in AD.

Using the results of the gene-based analyses described above, we tested whether the receptor complex and the two pathway branches contained significantly more AD associated genes as compared with the rest of the genome. We found that they were significantly (or almost significantly) enriched for genes associated to AD in the *APOE-*ε4 homozygotes (p-values 0.06, 0.009, 0.075, for the receptor complex and branches 1 and 2, respectively). The strongest significance was achieved when we combined the receptor complex and the two branches of the pathway (p=0.0055). Table 2 shows the details for each gene.

**Table 2.** Results of the gene-based analyses for the genes in the *DAB1-RELN* pathway accounting for the number of SNPs and the LD structure for each gene using MAGMA software.

| **GENE** | **CHR** | **START** | **STOP** | **N SNPs** | **p-value** |
| --- | --- | --- | --- | --- | --- |
| AKT1 | 14 | 105235686 | 105262080 | 76 | 0.0044 |
| CDH2 | 18 | 25530930 | 25757445 | 471 | 0.164 |
| CRK | 17 | 1324647 | 1359561 | 90 | 0.660 |
| CRKL | 22 | 21271714 | 21308037 | 86 | 0.847 |
| CTNND1 | 11 | 57520756 | 57586652 | 86 | 0.157 |
| DAB1 | 1 | 57460453 | 58716211 | 3496 | 0.004 |
| FYN | 6 | 111981535 | 112194655 | 538 | 0.456 |
| RAPGEF1 | 9 | 134452157 | 134615364 | 393 | 0.404 |
| GSK3B | 3 | 119540800 | 119813264 | 407 | 0.840 |
| PDK1 | 2 | 173420101 | 173490351 | 200 | 0.546 |
| PDK2 | 17 | 48172101 | 48188733 | 53 | 0.143 |
| PIK3CA | 3 | 178866311 | 178952500 | 203 | 0.033 |
| RELN | 7 | 103112231 | 103629963 | 1987 | 0.0069 |
| RAP1A | 1 | 112162405 | 112256807 | 307 | 0.00025 |
| SRC | 20 | 35973088 | 36033835 | 133 | 0.546 |
| VLDLR | 9 | 2621679 | 2654485 | 103 | 0.158 |
| YES1 | 18 | 721592 | 812327 | 232 | 0.809 |
| LRP8 | 1 | 53708036 | 53793821 | 194 | 0.842 |
| STK25 | 2 | 242434122 | 242449145 | 45 | 0.161 |

GENE –gene annotation with assembly hg19; CHR- chromosome; START, STOP – start and stop base-pair positions for genes; N SNPs- the number of SNPs in the analysis, p-value for the gene-based association test.

## Discussion

### No residual association at the APOE locus

*APOE*-ε4 is the strongest genetic risk factor for late onset AD. *APOE*-ε4 carriers have elevated risk for AD and earlier age-at-onset, with *APOE-*ε4 homozygotes at the highest risk [37, 38]. Many loci beyond *APOE* have been reported as associated with disease in increasingly large GWAS and meta-analyses, with over 80 susceptibility loci reported collectively [4, 39, 40]. We find no evidence to support the role of additional loci in an extended 2Mb region around *APOE* in *APOE-*ε4 homozygotes. This is supported by previous work on risk in the *APOE* region after adjusting for number of ε4 alleles [41, 42]. It is therefore unlikely that variants contribute additional risk to AD in the *APOE* region in *APOE*-ε4 homozygotes although association has previously been reported in *PVRL2* and *APOC1* in Chinese samples after adjusting for number of *APOE-*ε4 alleles [43]. Variants around *APOE* may explain additional variation in risk in populations where polymorphisms are in less pronounced LD with rs429358, and residual variability in *APOE*-ε3 carriers may still modify risk for the disease [44].

### Other established GWAS hits

This study does not have statistical power to reliably determine whether all the previously reported GWAS hits are associated with disease in *APOE-*ε4 homozygotes or whether those which do show direct evidence for association (nominal significance) are grouped in any particular pathway.

### Association with DAB1

Putative novel risk SNPs with strong evidence for association were mapped to the *DAB1* gene on chromosome 1. Roles for *DAB1* and *RELN* have previously been suggested in AD primarily based on studies in mice [36, 45–48] and functional genomic analysis in humans [49], but genome-wide association in humans has been lacking. However, it has been shown that the expression of *DAB1* and *RELN* are altered in AD brains [50–52]. DAB1 interacts with Asp-Pro-any residue-Tyr (NPXY) motifs in the cytoplasmic domains of amyloid precursor protein (APP) as it does with similar motifs in the cytoplasmic tails of the Reelin receptors through its N-terminal PTB domain [53, 54]. The NPXY motif is required for APP internalization and its deletion reduces Aβ production [55]. DAB1 association with APP has been shown to reduce amyloidogenic processing [36], which suggests it is involved in the intracellular trafficking of APP. Reelin also reduces Aβ production in HEK293 cells that don’t express DAB1 [47]. In a mouse model of AD, heterozygosity of *Reln* increases the accumulation of Aβ plaques [45], suggesting that the pathway physiologically alters APP cleavage in a manner that would protect against AD. In addition, homozygous loss-of-function in *Reln* and *Dab1* have been shown to augment tau-phosphorylation [21]. Reelin overexpression reduces abnormal somatodendritic localization of phosphor-Tau, Aβ plaques and synaptic loss in AD model mice [46, 48]. Thus there are links between the Reelin-DAB1 pathway and the two major pathological features of AD. In this study, both examined branches of the *DAB1-RELN* pathway had genes with significant association with AD. SNPs near *RAP1A* were significant; however, it remains to be determined if this branch regulates Aβ phosphor-Tau or another AD related pathology. The other major pathway downstream of Reelin-DAB1 has been associated with tau-phosphorylation and both *AKT* and *PIK3KA* from this branch were significantly associated with AD.

The dependence of the association between *DAB1*/*RELN* and AD on *APOE-*ε4 homozygosity is intriguing since there are several links between the Reelin pathway and APOE. The Reelin receptors are also APOE receptors and DAB1 binds the NPXY motifs in the cytoplasmic tails of other LDL-superfamily receptors [53, 54, 56], such as LDL-receptor related protein 1 that has roles in APOE/Aβ internalization and clearance [57]. Recent studies show that *APOE-*ε*4* reduces recycling of ApoER2 back to the plasma membrane making the cells less responsive to Reelin [58] and that Reelin protects against the toxic effects of Aβ on synapses [59]. Thus in *APOE*-ε4 homozygotes, one can imagine a threshold effect with high *APOE*-ε4 driving a pathological cycle by reducing the effects of DAB1 and RELN signalling including its normal function to reduce Aβ production/toxicity and/or MAPT-phosphorylation.

While the effect the SNPs have on the function of *DAB1* or other pathway genes remain to be determined, based on previous studies it would seem likely that they cause a partial loss-of-function that is potentially age dependent or cell-type specific in nature and would result in altered expression (eQTL) or splicing (sQTL). More than partial disruption of activity would likely lead to a developmental disorder in the homozygous individuals similar to loss-of-function alleles for *Dab1* in mice and *RELN* in humans and mice [60]. The significant SNPs identified here fall in intron 2 and are found in 4-7% of the population. Interestingly *DAB1* exomic variation is constrained and few variants are more prevalent than 1-2% (GnomAD) suggesting that the identified SNPs do not flag an alteration in the *DAB1* coding sequence. *DAB1* is alternatively spliced and differentially expressed most notably in a cell-type specific manner [26, 61–63]. Alternative splicing has been shown to regulate exons encoding a subset of the phosphorylation sites and a C-terminal exon altering *Dab1* functionality in mice. We note that humans have a read through variant of exon 3 that would lead to transcriptional termination 14 residues later (variant 9) that has not been identified in mice. It encodes the first part of the phosphotyrosine binding (PTB) domain residues 37-69, but it is likely to be functionally inert since the PTB domain extends to residue 171 [64]. With this complexity and the size of the *DAB1* gene, over 1 Mb, it could take significant effort to dissect the consequence of the SNPs identified here on gene function and AD.

In conclusion, we find a novel genome-wide significant hit in *DAB1* in an *APOE-*ε4 homozygote AD GWAS. This seems to be a hit only in *APOE-*ε4 homozygotes. Furthermore, it seems that this association marks a more general importance of the *DAB1-RELN* pathway in disease pathogenesis. It is not clear why this pathway should be of such importance in *APOE-*ε4 homozygotes only, but a clue may be that such individuals have particularly dense Aβ pathology and one can imagine that this pathway either has a role in modulating APP processing or in driving tau-phosphorylation in a manner that is dependent on high Aβ levels. This work suggests that *DAB1* has a protective role in late onset AD and highlights the importance of resolving the mechanism that likely involves the REELIN-DAB1 pathway for therapeutic development.

## Data Availability

Data for this study was obtained under UK Biobank application 15175. Data is available from the UK Biobank at www.ukbiobank.ac.uk.

## References

1. Hardy J, Escott-Price V. Genes, pathways and risk prediction in Alzheimer’s disease. Hum Mol Genet 2019; 28: R235–R240.

2. Coon KD, Myers AJ, Craig DW, Webster JA, Pearson J V., Lince DH, et al. A High-Density Whole-Genome Association Study Reveals That APOE Is the Major Susceptibility Gene for Sporadic Late-Onset Alzheimer’s Disease. J Clin Psychiatry 2007; 68: 0–0.

3. Leonenko G, Shoai M, Bellou E, Sims R, Williams J, Hardy J, et al. Genetic risk for alzheimer disease is distinct from genetic risk for amyloid deposition. Ann Neurol 2019; 86: 427–435.

4. Wightman DP, Jansen IE, Savage JE, Shadrin AA, Bahrami S, Holland D, et al. A genome-wide association study with 1,126,563 individuals identifies new risk loci for Alzheimer’s disease. Nat Genet 2021 539 2021; 53: 1276–1282.

5. Sudlow C, Gallacher J, Allen N, Beral V, Burton P, Danesh J, et al. UK Biobank: An Open Access Resource for Identifying the Causes of a Wide Range of Complex Diseases of Middle and Old Age. PLOS Med 2015; 12: e1001779.

6. Bycroft C, Freeman C, Petkova D, Band G, Elliott LT, Sharp K, et al. The UK Biobank resource with deep phenotyping and genomic data. Nature 2018; 562: 203–209.

7. Chang CC, Chow CC, Tellier LC, Vattikuti S, Purcell SM, Lee JJ. Second-generation PLINK: rising to the challenge of larger and richer datasets. Gigascience 2015; 4: 7.

8. Boughton AP, Welch RP, Flickinger M, VandeHaar P, Taliun D, Abecasis GR, et al. LocusZoom.js: interactive and embeddable visualization of genetic association study results. Bioinformatics 2021; 37: 3017–3018.

9. Hunter JD. Matplotlib. Comput Sci Eng 2007; 9: 90–95.

10. Leeuw CA de, Mooij JM, Heskes T, Posthuma D. MAGMA: Generalized Gene-Set Analysis of GWAS Data. PLoS Comput Biol 2015; 11.

11. Watanabe K, Taskesen E, van Bochoven A, Posthuma D. Functional mapping and annotation of genetic associations with FUMA. Nat Commun 2017 81 2017; 8: 1–11.

12. Lee GH, D’Arcangelo G. New insights into reelin-mediated signaling pathways. Front Cell Neurosci 2016; 10: 122.

13. Arnaud L, Ballif BA, Förster E, Cooper JA. Fyn Tyrosine Kinase Is a Critical Regulator of Disabled-1 during Brain Development. Curr Biol 2003; 13: 9–17.

14. Bock HH, Herz J. Reelin Activates Src Family Tyrosine Kinases in Neurons. Curr Biol 2003; 13: 18–26.

15. D’Arcangelo G, Homayouni R, Keshvara L, Rice DS, Sheldon M, Curran T. Reelin Is a Ligand for Lipoprotein Receptors. Neuron 1999; 24: 471–479.

16. Hiesberger T, Trommsdorff M, Howell BW, Goffinet A, Mumby MC, Cooper JA, et al. Direct Binding of Reelin to VLDL Receptor and ApoE Receptor 2 Induces Tyrosine Phosphorylation of Disabled-1 and Modulates Tau Phosphorylation. Neuron 1999; 24: 481–489.

17. Ballif BA, Arnaud L, Arthur WT, Guris D, Imamoto A, Cooper JA. Activation of a Dab1/CrkL/C3G/Rap1 Pathway in Reelin-Stimulated Neurons. Curr Biol 2004; 14: 606–610.

18. Franco SJ, Martinez-Garay I, Gil-Sanz C, Harkins-Perry SR, Müller U. Reelin Regulates Cadherin Function via Dab1/Rap1 to Control Neuronal Migration and Lamination in the Neocortex. Neuron 2011; 69: 482–497.

19. Jossin Y, Cooper JA. Reelin, Rap1 and N-cadherin orient the migration of multipolar neurons in the developing neocortex. Nat Neurosci 2011 146 2011; 14: 697–703.

20. Bock HH, Jossin Y, Liu P, Förster E, May P, Goffinet AM, et al. Phosphatidylinositol 3-Kinase Interacts with the Adaptor Protein Dab1 in Response to Reelin Signaling and Is Required for Normal Cortical Lamination *. J Biol Chem 2003; 278: 38772–38779.

21. Brich J, Shie FS, Howell BW, Li R, Tus K, Wakeland EK, et al. Genetic Modulation of Tau Phosphorylation in the Mouse. J Neurosci 2003; 23: 187–192.

22. Matsuki T, Zaka M, Guerreiro R, van der Brug MP, Cooper JA, Cookson MR, et al. Identification of Stk25 as a Genetic Modifier of Tau Phosphorylation in Dab1-Mutant Mice. PLoS One 2012; 7: e31152.

23. Escott-Price V, Shoai M, Pither R, Williams J, Hardy J. Polygenic score prediction captures nearly all common genetic risk for Alzheimer’s disease. Neurobiol Aging 2017; 49: 214.e7-214.e11.

24. Kunkle BW, Grenier-Boley B, Sims R, Bis JC, Damotte V, Naj AC, et al. Genetic meta-analysis of diagnosed Alzheimer’s disease identifies new risk loci and implicates Aβ, tau, immunity and lipid processing. Nat Genet 2019 513 2019; 51: 414–430.

25. Frieden C, Garai K. Structural differences between apoE3 and apoE4 may be useful in developing therapeutic agents for Alzheimer’s disease. Proc Natl Acad Sci U S A 2012; 109: 8913–8918.

26. Abadesco AD, Cilluffo M, Yvone GM, Carpenter EM, Howell BW, Phelps PE. Novel Disabled-1-expressing neurons identified in adult brain and spinal cord. Eur J Neurosci 2014; 39: 579–592.

27. Howell BW, Herrick TM, Cooper JA. Reelin-induced tryosine phosphorylation of Disabled 1 during neuronal positioning. Genes Dev 1999; 13: 643.

28. Howell BW, Herrick TM, Hildebrand JD, Zhang Y, Cooper JA. Dab1 tyrosine phosphorylation sites relay positional signals during mouse brain development. Curr Biol 2000; 10: 877–885.

29. Rice DS, Sheldon M, D’Arcangelo G, Nakajima K, Goldowitz D, Curran T. Disabled-1 acts downstream of Reelin in a signaling pathway that controls laminar organization in the mammalian brain. Development 1998; 125: 3719–3729.

30. Qiu S, Weeber EJ. Reelin signaling facilitates maturation of CA1 glutamatergic synapses. J Neurophysiol 2007; 97: 2312–2321.

31. Ventruti A, Kazdoba TM, Niu S, D’Arcangelo G. Reelin deficiency causes specific defects in the molecular composition of the synapses in the adult brain. Neuroscience 2011; 189: 32–42.

32. Pujadas L, Gruart A, Bosch C, Delgado L, Teixeira CM, Rossi D, et al. Reelin Regulates Postnatal Neurogenesis and Enhances Spine Hypertrophy and Long-Term Potentiation. J Neurosci 2010; 30: 4636–4649.

33. Rogers JT, Rusiana I, Trotter J, Zhao L, Donaldson E, Pak DTS, et al. Reelin supplementation enhances cognitive ability, synaptic plasticity, and dendritic spine density. Learn Mem 2011; 18: 558–564.

34. Trotter J, Lee GH, Kazdoba TM, Crowell B, Domogauer J, Mahoney HM, et al. Dab1 Is Required for Synaptic Plasticity and Associative Learning. J Neurosci 2013; 33: 15652–15668.

35. Weeber EJ, Beffert U, Jones C, Christian JM, Förster E, David Sweatt J, et al. Reelin and ApoE Receptors Cooperate to Enhance Hippocampal Synaptic Plasticity and Learning *. J Biol Chem 2002; 277: 39944–39952.

36. Hoe H-S, Tran TS, Matsuoka Y, Howell BW, Rebeck GW. DAB1 and Reelin Effects on Amyloid Precursor Protein and ApoE Receptor 2 Trafficking and Processing *. J Biol Chem 2006; 281: 35176–35185.

37. Corder EH, Saunders AM, Strittmatter WJ, Schmechel DE, Gaskell PC, Small GW, et al. Gene Dose of Apolipoprotein E Type 4 Allele and the Risk of Alzheimer’s Disease in Late Onset Families. Science (80-) 1993; 261: 921–923.

38. Freudenberg-Hua Y, Li W, Davies P. Effects of Age, Sex, and Ethnicity on the Association Between Apolipoprotein E Genotype and Alzheimer Disease: A Meta-analysis. Front Med 2018; 0: 108.

39. Andrews SJ, Fulton-Howard B, Goate A. Interpretation of risk loci from genome-wide association studies of Alzheimer’s disease. Lancet Neurol 2020; 19: 326–335.

40. Bellenguez C, Küçükali F, Jansen IE, Kleineidam L, Moreno-Grau S, Amin N, et al. New insights into the genetic etiology of Alzheimer’s disease and related dementias. Nat Genet 2022 544 2022; 54: 412–436.

41. Jun G, Vardarajan BN, Buros J, Yu C-E, Hawk M V., Dombroski BA, et al. Comprehensive Search for Alzheimer Disease Susceptibility Loci in the APOE Region. Arch Neurol 2012; 69: 1270–1279.

42. Naj AC, Jun G, Beecham GW, Wang L-S, Vardarajan BN, Buros J, et al. Common variants at MS4A4/MS4A6E, CD2AP, CD33 and EPHA1 are associated with late-onset Alzheimer’s disease. Nat Genet 2011 435 2011; 43: 436–441.

43. Zhou X, Chen Y, Mok KY, Kwok TCY, Mok VCT, Guo Q, et al. Non-coding variability at the APOE locus contributes to the Alzheimer’s risk. Nat Commun 2019 101 2019; 10: 1–16.

44. Roses AD, Lutz MW, Amrine-Madsen H, Saunders AM, Crenshaw DG, Sundseth SS, et al. A TOMM40 variable-length polymorphism predicts the age of late-onset Alzheimer’s disease. Pharmacogenomics J 2010 105 2009; 10: 375–384.

45. Kocherhans S, Madhusudan A, Doehner J, Breu KS, Nitsch RM, Fritschy JM, et al. Reduced Reelin Expression Accelerates Amyloid-β Plaque Formation and Tau Pathology in Transgenic Alzheimer’s Disease Mice. J Neurosci 2010; 30: 9228–9240.

46. Pujadas L, Rossi D, Andrés R, Teixeira CM, Serra-Vidal B, Parcerisas A, et al. Reelin delays amyloid-beta fibril formation and rescues cognitive deficits in a model of Alzheimer’s disease. Nat Commun 2014 51 2014; 5: 1–11.

47. Rice HC, Young-Pearse TL, Selkoe DJ. Systematic evaluation of candidate ligands regulating ectodomain shedding of Amyloid precursor protein. Biochemistry 2013; 52: 3264–3277.

48. Rossi D, Gruart A, Contreras-Murillo G, Muhaisen A, Ávila J, Delgado-García JM, et al. Reelin reverts biochemical, physiological and cognitive alterations in mouse models of Tauopathy. Prog Neurobiol 2020; 186: 101743.

49. Gao H, Tao Y, He Q, Song F, Saffen D. Functional enrichment analysis of three Alzheimer’s disease genome-wide association studies identities DAB1 as a novel candidate liability/protective gene. Biochem Biophys Res Commun 2015; 463: 490–495.

50. Botella-López A, Burgaya F, Gavín R, García-Ayllón MS, Gómez-Tortosa E, Peña-Casanova J, et al. Reelin expression and glycosylation patterns are altered in Alzheimer’s disease. Proc Natl Acad Sci U S A 2006; 103: 5573–5578.

51. Muller T, Loosse C, Schrotter A, Schnabel A, Helling S, Egensperger R, et al. The AICD Interacting Protein DAB1 is Up-Regulated in Alzheimer Frontal Cortex Brain Samples and Causes Deregulation of Proteins Involved in Gene Expression Changes. Curr Alzheimer Res 2011; 8: 573–582.

52. Chin J, Massaro CM, Palop JJ, Thwin MT, Yu GQ, Bien-Ly N, et al. Reelin Depletion in the Entorhinal Cortex of Human Amyloid Precursor Protein Transgenic Mice and Humans with Alzheimer’s Disease. J Neurosci 2007; 27: 2727–2733.

53. Howell BW, Lanier LM, Frank R, Gertler FB, Cooper JA. The Disabled 1 Phosphotyrosine-Binding Domain Binds to the Internalization Signals of Transmembrane Glycoproteins and to Phospholipids. Mol Cell Biol 1999; 19: 5179–5188.

54. Trommsdorff M, Borg JP, Margolis B, Herz J. Interaction of Cytosolic Adaptor Proteins with Neuronal Apolipoprotein E Receptors and the Amyloid Precursor Protein *. J Biol Chem 1998; 273: 33556–33560.

55. Perez RG, Soriano S, Hayes JD, Ostaszewski B, Xia W, Selkoe DJ, et al. Mutagenesis Identifies New Signals for β-Amyloid Precursor Protein Endocytosis, Turnover, and the Generation of Secreted Fragments, Including Aβ42 *. J Biol Chem 1999; 274: 18851–18856.

56. Howell BW, Herz J. The LDL receptor gene family: signaling functions during development. Curr Opin Neurobiol 2001; 11: 74–81.

57. Shinohara M, Tachibana M, Kanekiyo T, Bu G. Role of LRP1 in the pathogenesis of Alzheimer’s disease: evidence from clinical and preclinical studies: Thematic Review Series: ApoE and Lipid Homeostasis in Alzheimer’s Disease. J Lipid Res 2017; 58: 1267–1281.

58. Chen Y, Durakoglugil MS, Xian X, Herz J. ApoE4 reduces glutamate receptor function and synaptic plasticity by selectively impairing ApoE receptor recycling. Proc Natl Acad Sci U S A 2010; 107: 12011–12016.

59. Lane-Donovan C, Philips GT, Wasser CR, Durakoglugil MS, Masiulis I, Upadhaya A, et al. Reelin protects against amyloid β toxicity in vivo. Sci Signal 2015; 8.

60. Bar I, Tissir F, Lambert de Rouvroit C, De Backer O, Goffinet AM. The Gene Encoding Disabled-1 (DAB1), the Intracellular Adaptor of the Reelin Pathway, Reveals Unusual Complexity in Human and Mouse *. J Biol Chem 2003; 278: 5802–5812.

61. D D, Hung K-Y, Tarn W-Y. RBM4 Modulates Radial Migration via Alternative Splicing of Dab1 during Cortex Development. Mol Cell Biol 2018; 38.

62. Gao Z, Godbout R. Reelin-Disabled-1 signaling in neuronal migration: splicing takes the stage. Cell Mol Life Sci 2012 7013 2012; 70: 2319–2329.

63. Yano M, Hayakawa-Yano Y, Mele A, Darnell RB. Nova2 Regulates Neuronal Migration through an RNA Switch in Disabled-1 Signaling. Neuron 2010; 66: 848–858.

64. Howell BW, Gertler FB, Cooper JA. Mouse disabled (mDab1): a Src binding protein implicated in neuronal development. EMBO J 1997; 16: 121–132.

65. Marioni RE, Harris SE, Zhang Q, McRae AF, Hagenaars SP, Hill WD, et al. GWAS on family history of Alzheimer’s disease. Transl Psychiatry 2018 81 2018; 8: 1–7.

