## Supplementary for "Whole genome analysis in *APOE4* homozygotes identifies the *DAB1-RELN* pathway in Alzheimer’s disease pathogenesis"

### Supplementary Information.

**Supplemental Table S1:** Previously reported genome-wide significant SNPs and their association results in the current study.

| | | | | *Current Analysis* | | | | | *Previous GWAS* | | | | |
|---|---|---|---|---|---|---|---|---|---|---|---|---|---|
| **CHR** | **BP** | **SNP** | **Closest gene** | **Effect/Alt** | **Freq** | **OR** | **SE** | ***p*-value** | **Effect/Alt** | **OR** | ***p*-value** | **GWAS** | ***power at 5% sig.level*** |
| 1 | 985377 | rs113020870 | *AGRN* | NA | NA | NA | NA | NA | C/T | 1.90 | $3.8x10^{-08}$ | Wi | NA |
| 1 | 109888432 | rs141749679 | SORT1 | NA | NA | NA | NA | NA | C/T | 1.38 | $7.5x10^{-09}$ | Be | NA |
| 1 | 161155392 | rs4575098 | *ADAMTS4* | A/G | 0.23 | 0.91 | 0.11 | $3.7x10^{-01}$ | A/G | 1.02 | $2.1x10^{-10}$ | Ja | 0.060 |
| 1 | 207802552 | rs4844610 | *CR1* | C/A | 0.82 | 0.84 | 0.11 | $1.2x10^{-01}$ | C/A | 0.85 | $3.6x10^{-24}$ | Ku | 0.105 |
| 2 | 9699011 | rs72777026 | ADAM17 | G/A | 0.14 | 0.87 | 0.13 | $2.7x10^{-01}$ | G/A | 1.06 | $2.7x10^{-08}$ | Be | 0.084 |
| 2 | 37531939 | rs17020490 | PRKD3 | C/T | 0.14 | 0.97 | 0.13 | $8.1x10^{-01}$ | C/T | 1.06 | $3.3x10^{-09}$ | Be | 0.084 |
| 2 | 106366056 | rs143080277 | NCK2 | NA | NA | NA | NA | NA | C/T | 1.47 | $2.1x10^{-13}$ | Be | NA |
| **2** | **127892810** | **rs6733839** | ***BIN1*** | **T/C** | **0.38** | **1.26** | **0.09** | $\mathbf{1.1x10^{-02}}$ | **T/C** | **1.2** | $\mathbf{2.4x10^{-69}}$ | **Ma** | **0.179** |

| | | | | | | | | | | | | | |
|---|---|---|---|---|---|---|---|---|---|---|---|---|---|
| 2 | 203743440 | rs139643391 | WDR12 | NA | NA | NA | NA | NA | T/TC | 0.94 | $1.1x10^{-08}$ | Be | NA |
| 2 | 234068476 | rs35349669 | *INPP5D* | T/C | 0.49 | 1.18 | 0.09 | $6.2x10^{-02}$ | T/C | 1.07 | $3.6x10^{-11}$ | Ma | 0.082 |
| 3 | 154787511 | rs16824536 | MME | A/G | 0.06 | 0.97 | 0.19 | $8.9x10^{-01}$ | A/G | 0.92 | $3.6x10^{-08}$ | Be | 0.099 |
| 3 | 154801978 | rs61762319 | MME | G/A | 0.03 | 1.40 | 0.23 | $1.3x10^{-01}$ | G/A | 1.16 | $2.2x10^{-11}$ | Be | 0.141 |
| 4 | 987343 | rs3822030 | IDUA | G/T | 0.43 | 1.06 | 0.09 | $5.2x10^{-01}$ | G/T | 0.95 | $8.3x10^{-12}$ | Be | 0.075 |
| 4 | 11026028 | rs6448453 | *CLNK* | G/A | 0.74 | 0.97 | 0.10 | $7.6x10^{-01}$ | G/A | 0.99 | $1.9x10^{-09}$ | Ja | 0.053 |
| 4 | 40198846 | rs2245466 | RHOH | G/C | 0.33 | 0.99 | 0.10 | $9.6x10^{-01}$ | G/C | 1.05 | $1.2x10^{-09}$ | Be | 0.075 |
| 5 | 14724413 | rs112403360 | ANKH | A/T | 0.08 | 1.25 | 0.15 | $1.5x10^{-01}$ | A/T | 1.09 | $2.3x10^{-09}$ | Be | 0.103 |
| 5 | 86223195 | rs62374257 | COX7C | C/T | 0.23 | 1.11 | 0.10 | $3.3x10^{-01}$ | C/T | 1.07 | $1.4x10^{-15}$ | Be | 0.090 |
| 5 | 88223420 | rs190982 | *MEF2C* | A/G | 0.60 | 1 | 0.09 | 1 | A/G | 1.08 | $3.2x10^{-08}$ | La | 0.082 |
| 5 | 150432388 | rs871269 | *TNIP1* | T/C | 0.32 | 0.98 | 0.09 | $8.0x10^{-01}$ | T/C | 0.79 | $1.4x10^{-09}$ | Wi | 0.273 |
| 5 | 156526331 | rs6891966 | *HAVCR2* | A/G | 0.23 | 0.87 | 0.11 | $2.1x10^{-01}$ | A/G | 0.86 | $7.9x10^{-10}$ | Wi | 0.169 |
| 5 | 179628150 | rs113706587 | RASGEF1C | A/G | 0.10 | 1.17 | 0.14 | $2.5x10^{-01}$ | A/G | 1.09 | $2.2x10^{-16}$ | Be | 0.105 |
| 6 | 32559825 | rs34855541 | *HLA-DRB1* | G/A | 0.19 | 0.88 | 0.12 | $2.5x10^{-01}$ | G/A | 0.9 | $9.5x10^{-15}$ | Ma | 0.123 |
| 6 | 47432637 | rs9381563 | *CD2AP* | T/C | 0.64 | 0.97 | 0.09 | $7.3x10^{-01}$ | T/C | 0.93 | $5.8x10^{-14}$ | Ma | 0.080 |
| 6 | 114612895 | rs785129 | HS3ST5 | T/C | 0.35 | 0.95 | 0.10 | $6.1x10^{-01}$ | T/C | 1.04 | $2.4x10^{-09}$ | Be | 0.069 |

| | | | | | | | | | | | | | |
|---|---|---|---|---|---|---|---|---|---|---|---|---|---|
| 7 | 7856894 | rs6943429 | UMAD1 | T/C | 0.41 | 1.10 | 0.09 | $3.1x10^{-01}$ | T/C | 1.05 | $1.0x10^{-10}$ | Be | 0.073 |
| 7 | 8244012 | rs10952097 | ICA1 | T/C | 0.11 | 0.97 | 0.14 | $8.1x10^{-01}$ | T/C | 1.07 | $6.8x10^{-09}$ | Be | 0.090 |
| 7 | 12269593 | rs13237518 | TMEM106B | A/C | 0.41 | 0.86 | 0.09 | $1.1x10^{-01}$ | A/C | 0.96 | $4.9x10^{-11}$ | Be | 0.069 |
| 7 | 28168750 | rs1160871 | JAZF1 | NA | NA | NA | NA | NA | G/GTCTT | 0.95 | $9.8x10^{-09}$ | Be | NA |
| 7 | 37841534 | rs2718058 | *GPR141* | G/A | 0.37 | 0.92 | 0.09 | $3.4x10^{-01}$ | G/A | 0.93 | $4.8x10^{-09}$ | La | 0.090 |
| **7** | **54941328** | **rs76928645** | **SEC61G** | **T/C** | **0.10** | **0.66** | **0.17** | **$1.5x10^{-02}$** | **T/C** | **0.93** | **$1.6x10^{-10}$** | **Be** | **0.094** |
| 7 | 100004446 | rs1476679 | *ZCWPW1* | T/C | 0.69 | 1.09 | 0.10 | $3.9x10^{-01}$ | T/C | 1.1 | $9.9x10^{-19}$ | Ma | 0.085 |
| 7 | 143099133 | rs10808026 | *EPHA1* | A/C | 0.22 | 0.93 | 0.11 | $4.8x10^{-01}$ | A/C | 0.91 | $1.1x10^{-14}$ | Ma | 0.112 |
| 8 | 11702122 | rs1065712 | CTSB | C/G | 0.05 | 1.14 | 0.20 | $5.1x10^{-01}$ | C/G | 1.09 | $1.9x10^{-09}$ | Be | 0.100 |
| 8 | 27464929 | rs4236673 | *CLU* | G/A | 0.60 | 1.17 | 0.09 | $8.7x10^{-02}$ | G/A | 1.12 | $1.1x10^{-28}$ | Ma | 0.101 |
| 8 | 145158607 | rs34173062 | SHARPIN | A/G | 0.07 | 0.93 | 0.19 | $7.2x10^{-01}$ | A/G | 1.13 | $1.7x10^{-16}$ | Be | 0.134 |
| 9 | 107665978 | rs1800978 | ABCA1 | G/C | 0.12 | 0.78 | 0.15 | $8.7x10^{-02}$ | G/C | 1.06 | $1.6x10^{-09}$ | Be | 0.084 |
| 10 | 11720308 | rs7920721 | *ECHDC3* | G/A | 0.38 | 1.02 | 0.09 | $8.3x10^{-01}$ | G/A | 1.08 | $1.8x10^{-11}$ | Ku | 0.091 |
| 10 | 61784928 | rs7068231 | ANK3 | T/G | 0.40 | 1.03 | 0.09 | $7.7x10^{-01}$ | T/G | 0.95 | $3.3x10^{-13}$ | Be | 0.076 |
| 10 | 82253984 | rs6586028 | TSPAN14 | C/T | 0.21 | 0.89 | 0.11 | $3.0x10^{-01}$ | C/T | 0.93 | $2.0x10^{-19}$ | Be | 0.094 |
| 10 | 98026407 | rs6584063 | BLNK | G/A | 0.04 | 0.70 | 0.26 | $1.6x10^{-01}$ | G/A | 0.89 | $6.7x10^{-11}$ | Be | 0.120 |

| | | | | | | | | | | | | | |
|---|---|---|---|---|---|---|---|---|---|---|---|---|---|
| 10 | 124172912 | rs7908662 | PLEKHA1 | G/A | 0.48 | 1.10 | 0.09 | $2.8x10^{-01}$ | G/A | 0.96 | $2.6x10^{-09}$ | Be | 0.068 |
| 11 | 47380340 | rs3740688 | *SPI1* | T/G | 0.55 | 1.1 | 0.09 | $3.0x10^{-01}$ | T/G | 1.09 | $5.5x10^{-13}$ | Ku | 0.089 |
| 11 | 59936926 | rs7933202 | *MS4A6A* | C/A | 0.40 | 0.86 | 0.09 | $8.8x10^{-02}$ | C/A | 0.89 | $1.9x10^{-19}$ | Ku | 0.122 |
| 11 | 85867875 | rs10792832 | *PICALM* | G/A | 0.63 | 1.04 | 0.09 | $7.0x10^{-01}$ | G/A | 1.13 | $5.1x10^{-36}$ | Ma | 0.102 |
| **11** | **121435587** | **rs11218343** | ***SORL1*** | **C/T** | **0.04** | **0.41** | **0.33** | $\mathbf{7.3x10^{-03}}$ | **C/T** | **0.81** | $\mathbf{4.6x10^{-17}}$ | **Ma** | **0.211** |
| 12 | 113719788 | rs6489896 | TPCN1 | C/T | 0.07 | 0.99 | 0.17 | $9.6x10^{-01}$ | C/T | 1.08 | $1.8x10^{-09}$ | Be | 0.096 |
| 14 | 53391680 | rs17125924 | *FERMT2* | G/A | 0.09 | 0.95 | 0.15 | $7.3x10^{-01}$ | G/A | 1.12 | $1.3x10^{-11}$ | Ma | 0.128 |
| 14 | 92938855 | rs12590654 | *SLC24A4* | A/G | 0.34 | 0.89 | 0.10 | $2.2x10^{-01}$ | A/G | 0.92 | $8.2x10^{-12}$ | Ma | 0.099 |
| 14 | 106228095 | rs7157106 | IGH gene cluster | A/G | 0.35 | 0.99 | 0.09 | $9.4x10^{-01}$ | A/G | 1.05 | $2.0x10^{-08}$ | Be | 0.075 |
| 14 | 107121607 | rs10131280 | IGH gene cluster | A/G | 0.13 | 0.84 | 0.14 | $2.0x10^{-01}$ | A/G | 0.94 | $4.3x10^{-10}$ | Be | 0.087 |
| 15 | 51001534 | rs59685680 | *SPPL2A* | G/T | 0.20 | 0.94 | 0.11 | $5.8x10^{-01}$ | G/T | 0.93 | $9.2x10^{-09}$ | Ma | 0.095 |
| 15 | 59045774 | rs593742 | *ADAM10* | G/A | 0.31 | 0.95 | 0.10 | $6.3x10^{-01}$ | G/A | 0.93 | $2.8x10^{-11}$ | Ma | 0.092 |
| **15** | **63569902** | **rs117618017** | ***APH1B*** | **T/C** | **0.15** | **1.3** | **0.12** | $\mathbf{2.4x10^{-02}}$ | **T/C** | **1.02** | $\mathbf{3.3x10^{-08}}$ | **Ja** | **0.060** |
| **15** | **64423506** | **rs3848143** | **SNX1** | **G/A** | **0.20** | **1.23** | **0.10** | $\mathbf{4.8x10^{-02}}$ | **G/A** | **1.05** | $\mathbf{8.4x10^{-11}}$ | **Be** | **0.077** |

| | | | | | | | | | | | | | |
|---|---|---|---|---|---|---|---|---|---|---|---|---|---|
| 15 | 79229199 | rs12592898 | CTSH | A/G | 0.13 | 0.94 | 0.13 | $6.1x10^{-01}$ | A/G | 0.94 | $4.2x10^{-09}$ | Be | 0.087 |
| **16** | **19808163** | **rs7185636** | ***IQCK*** | **C/T** | **0.17** | **0.72** | **0.13** | **$9.0x10^{-03}$** | **C/T** | **0.92** | **$2.4x10^{-08}$** | **Ku** | **0.103** |
| 16 | 30021402 | rs1140239 | DOC2A | NA | NA | NA | NA | NA | T/C | 0.94 | $2.6x10^{-13}$ | Be | NA |
| 16 | 31133100 | rs59735493 | *KAT8* | A/G | 0.28 | 1.01 | 0.10 | $9.0x10^{-01}$ | A/G | 0.99 | $4.0x10^{-08}$ | Ja | 0.055 |
| 16 | 70694000 | rs4985556 | *IL34* | A/C | 0.11 | 0.98 | 0.14 | $8.7x10^{-01}$ | A/C | 1.09 | $3.7x10^{-08}$ | Ma | 0.105 |
| 16 | 79355857 | rs62039712 | *WWOX* | A/G | 0.12 | 0.88 | 0.14 | $3.8x10^{-01}$ | A/G | 1.16 | $3.7x10^{-08}$ | Ku | 0.166 |
| 16 | 79608408 | rs450674 | MAF | C/T | 0.40 | 0.87 | 0.09 | $1.3x10^{-01}$ | C/T | 0.96 | $3.2x10^{-08}$ | Be | 0.070 |
| 16 | 81773209 | rs12444183 | *PLCG2* | G/A | 0.61 | 0.98 | 0.09 | $8.5x10^{-01}$ | G/A | 1.06 | $3.2x10^{-08}$ | Ma | 0.073 |
| 16 | 86454210 | rs16941239 | FOXF1 | A/T | 0.02 | 1.56 | 0.25 | $7.5x10^{-02}$ | A/T | 1.13 | $1.3x10^{-08}$ | Be | 0.113 |
| 16 | 90170095 | rs56407236 | PRDM7 | A/G | 0.07 | 0.91 | 0.18 | $5.8x10^{-01}$ | A/G | 1.11 | $6.5x10^{-15}$ | Be | 0.118 |
| 17 | 1631350 | rs35048651 | WDR81 | NA | NA | NA | NA | NA | T/TGAG | 1.06 | $7.7x10^{-11}$ | Be | NA |
| 17 | 5137047 | rs7225151 | *SCIMP* | A/G | 0.12 | 1 | 0.14 | $9.8x10^{-01}$ | A/G | 1.1 | $6.1x10^{-12}$ | Ma | 0.113 |
| 17 | 18059454 | rs2242595 | MYO15A | A/G | 0.12 | 0.81 | 0.14 | $1.5x10^{-01}$ | A/G | 0.94 | $1.1x10^{-09}$ | Be | 0.087 |
| 17 | 42430244 | rs5848 | GRN | T/C | 0.27 | 1.02 | 0.10 | $8.7x10^{-01}$ | T/C | 1.07 | $2.4x10^{-20}$ | Be | 0.088 |
| 17 | 42442344 | rs708382 | *GRN* | C/T | 0.39 | 1.02 | 0.09 | $7.9x10^{-01}$ | C/T | 1.31 | $2.0x10^{-09}$ | Wi | 0.275 |
| 17 | 47450775 | rs28394864 | *RP11-81K2.1* | A/G | 0.46 | 1.07 | 0.09 | $4.3x10^{-01}$ | A/G | 1.01 | $1.9x10^{-08}$ | Ja | 0.054 |

| | | | | | | | | | | | | | |
|---|---|---|---|---|---|---|---|---|---|---|---|---|---|
| 17 | 56398006 | rs2526380 | *BZRAP1* | G/C | 0.43 | 1.04 | 0.09 | $6.4x10^{-01}$ | G/C | 0.97 | $2.6x10^{-08}$ | Ja | 0.064 |
| 17 | 61538148 | rs138190086 | *CYB561* | A/G | 0.02 | 0.99 | 0.33 | $9.7x10^{-01}$ | A/G | 1.25 | $1.9x10^{-09}$ | Ma | 0.187 |
| 18 | 56189459 | rs76726049 | *ALPK2* | C/T | 0.02 | 0.9 | 0.38 | $7.8x10^{-01}$ | C/T | 1.06 | $3.3x10^{-08}$ | Ja | 0.072 |
| 19 | 1056492 | rs3752246 | *ABCA7* | C/G | 0.83 | 0.92 | 0.11 | $4.3x10^{-01}$ | C/G | 0.87 | $3.1x10^{-16}$ | Ku | 0.094 |
| 19 | 1854254 | rs149080927 | KLF16 | NA | NA | NA | NA | NA | G/GC | 1.05 | $5.1x10^{-10}$ | Be | NA |
| 19 | 45411941 | rs429358 | *APOE* | NA | NA | NA | NA | NA | T/C | 3.32 | $1.2x10^{-881}$ | Ku | NA |
| 19 | 49213504 | rs2452170 | *NTN5* | A/G | 0.53 | 1.16 | 0.09 | $1.0x10^{-01}$ | A/G | 1.24 | $1.7x10^{-08}$ | Wi | 0.181 |
| 19 | 50453317 | rs9304690 | SIGLEC11 | T/C | 0.25 | 1.08 | 0.10 | $4.2x10^{-01}$ | T/C | 1.05 | $4.7x10^{-09}$ | Be | 0.076 |
| **19** | **51728477** | **rs12459419** | ***CD33*** | **T/C** | **0.33** | **0.75** | **0.10** | $\mathbf{3.5x10^{-03}}$ | **T/C** | **0.99** | $\mathbf{6.3x10^{-09}}$ | **Ma** | **0.055** |
| **19** | **54771451** | **rs587709** | **LILRB2** | **C/T** | **0.27** | **1.31** | **0.10** | $\mathbf{5.1x10^{-03}}$ | **C/T** | **1.05** | $\mathbf{3.6x10^{-11}}$ | **Be** | **0.076** |
| 19 | 54825174 | rs1761461 | *LILRB2* | C/A | 0.50 | 0.84 | 0.09 | $5.2x10^{-02}$ | C/A | 0.84 | $1.6x10^{-09}$ | Wi | 0.166 |
| 20 | 393978 | rs1358782 | RBCK1 | A/G | 0.23 | 1.04 | 0.10 | $7.3x10^{-01}$ | A/G | 0.95 | $1.6x10^{-08}$ | Be | 0.079 |
| 20 | 54983075 | rs6069736 | *CSTF1* | T/C | 0.09 | 0.81 | 0.17 | $2.0x10^{-01}$ | T/C | 0.89 | $2.0x10^{-10}$ | Ma | 0.132 |
| 20 | 62374441 | rs6742 | SLC2A4RG | T/C | 0.22 | 1.00 | 0.11 | $9.8x10^{-01}$ | T/C | 0.95 | $2.6x10^{-09}$ | Be | 0.079 |
| 21 | 27473875 | rs2154481 | APP | C/T | 0.48 | 0.89 | 0.09 | $1.9x10^{-01}$ | C/T | 0.95 | $1.0x10^{-12}$ | Be | 0.074 |

| | | | | | | | | | | | | | |
|---|---|---|---|---|---|---|---|---|---|---|---|---|---|
| 21 | 28156856 | rs2830500 | *ADAMTS1* | A/C | 0.29 | 0.84 | 0.10 | $9.2x10^{-02}$ | A/C | 0.93 | $2.6x10^{-08}$ | Ku | 0.092 |

CHR –chromosome; BP –base-pair position in build37; SNP –single nucleotide polymorphism, closest gene –genes were annotated with assembly hg19; effect/non-effect –effect and non-effect alleles; freq- frequency of reference allele in the UK Biobank *APOE*-ε4 homozygotes individuals; OR, SE, p-value –odds ratio, standard error and p-value of the current and previous reported AD GWAS association studies; GWAS –corresponding GWAS study (Ma = Marioni et al., Ku = Kunkle et al., Ja = Jansen et al., La = Lambert et al., Wi = Wightman et al., Be = Bellenguez et al 2021); power at 5% sig.level - power to detect the reported effect size in ε4ε4 homozygous sample of UK Biobank at 5% significance level. No proxy SNP with $R^2$ > 0.7 was found for rs184384746 (closest gene *HESX1),* rs187370608 (*UNC5CL)* or rs114360492 (*CNTNAP2)*. Starred SNPs were present on the genotyping array (rather than imputed). Replicated loci are shown in bold.

**Supplemental Table S2**. Allele frequencies of GWAS significant SNPs in DAB1, depending upon AD case-control status and APOE-ε4 genotype.

| N *APOE*-ε4 alleles | AD status | rs17541203_C | rs197111_T | rs78921149_T | rs112437613_T | rs17115257_G | rs58359668_T |
|---|---|---|---|---|---|---|---|
| 0 | 0 | 0.067 | 0.069 | 0.066 | 0.066 | 0.081 | 0.081 |
| | 1 | 0.066 | 0.066 | 0.064 | 0.064 | 0.081 | 0.081 |
| 1 | 0 | 0.066 | 0.068 | 0.065 | 0.065 | 0.080 | 0.08 |
| | 1 | 0.069 | 0.072 | 0.069 | 0.069 | 0.086 | 0.086 |
| 2 | 0 | 0.063 | 0.065 | 0.062 | 0.061 | 0.076 | 0.076 |
| | 1 | 0.12 | 0.12 | 0.12 | 0.12 | 0.14 | 0.14 |

N *APOE*-ε4 alleles –number of *APOE*- ε4 alleles (coded 0,1,2); AD –Alzheimer's disease; AD status – coded AD case -1 and control -0; rs-number _SNP rs number and corresponding minor allele.

**Figure S1.** LocusZoom plot showing SNP associations in the *APOE* region (chromosome 19: 44.5-46.5 Mb).

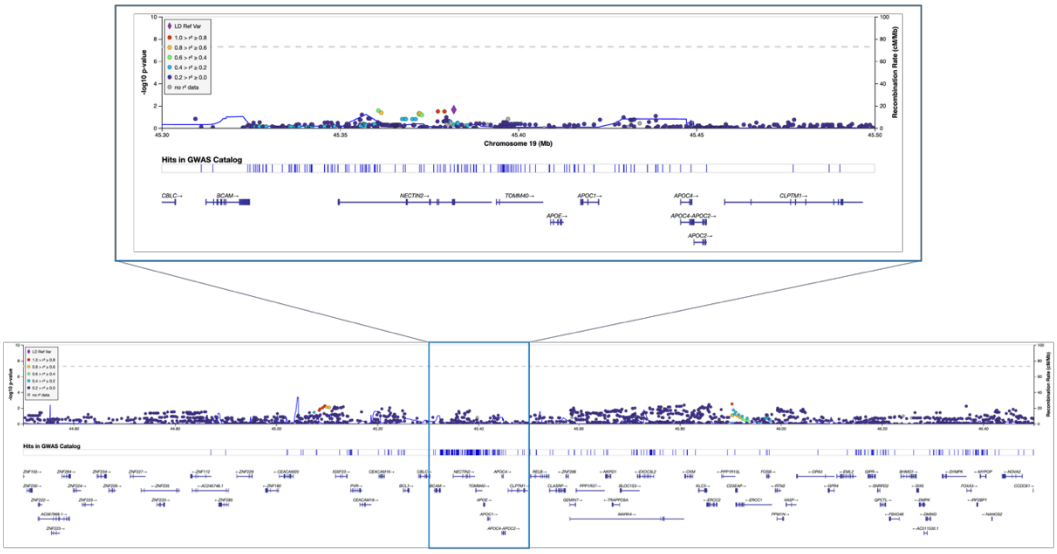

**Figure S2.** LocusZoom plot showing SNP associations in the *DAB1* gene*.*

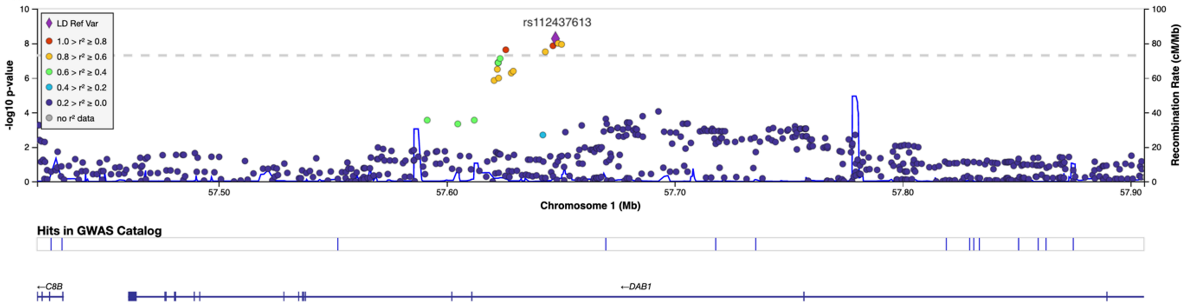

**Figure S3**: LocusZoom plot showing SNP associations in the *RELN* gene.

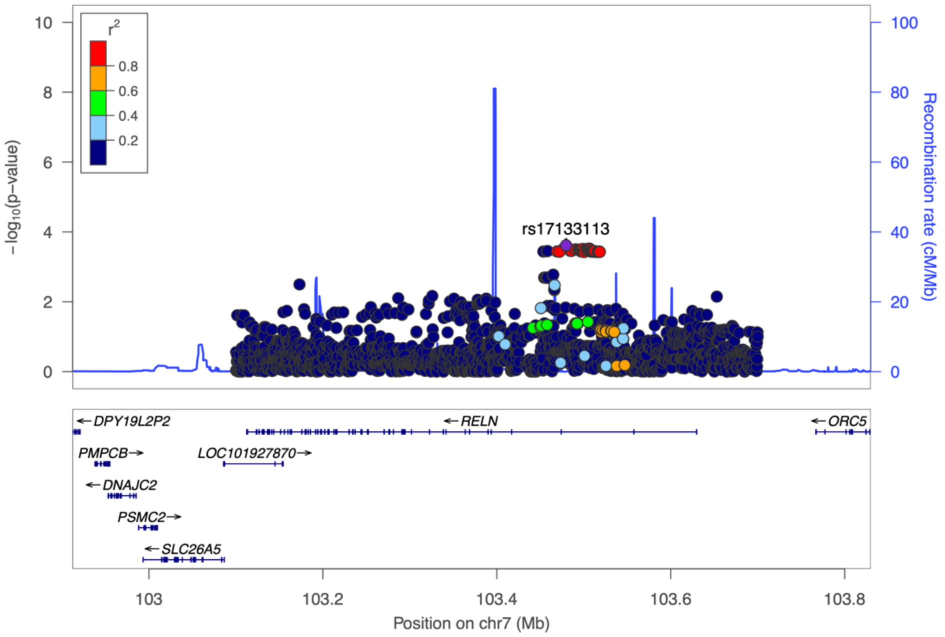

**Figure S4**: Epistatic effect between *APOE*-ε4 and rs17133113 (*RELN*) in the whole sample of the UK Biobank aged 65+ (N=229,748).

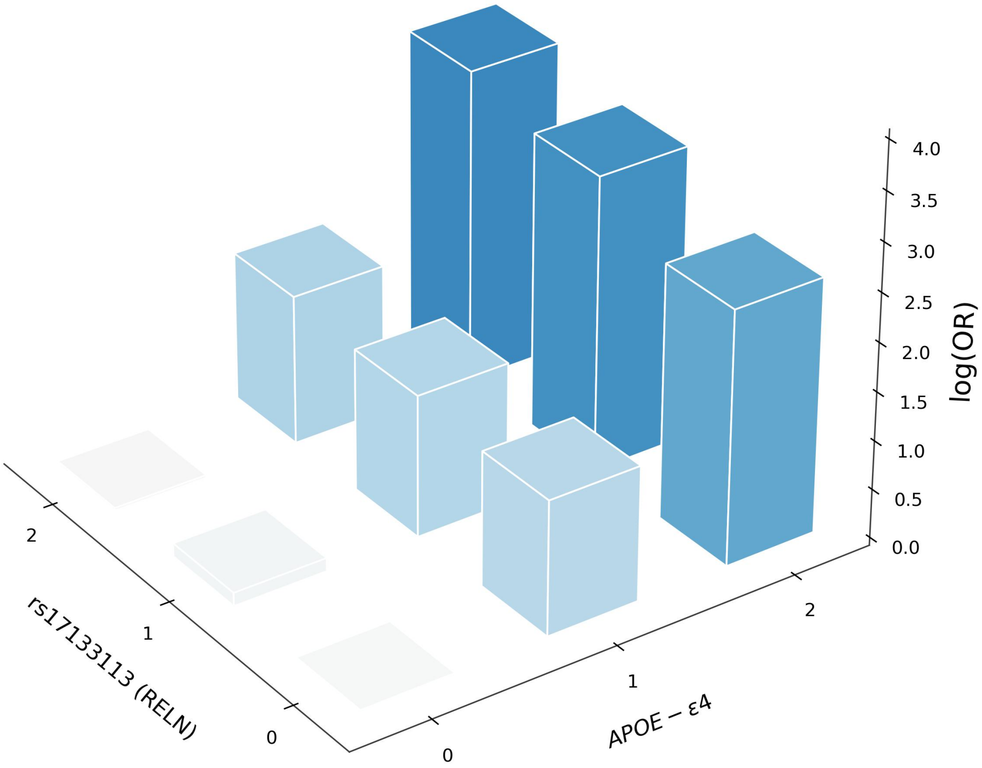
